# Reduction in COPD exacerbations during COVID-19: a systematic review and meta-analysis

**DOI:** 10.1101/2021.05.17.21257335

**Authors:** Jaber S. Alqahtani, Tope Oyelade, Abdulelah M. Aldhahir, Renata Gonçalves Mendes, Saeed M. Alghamdi, Marc Miravitlles, Swapna Mandal, John R. Hurst

## Abstract

**Background:** Reports have suggested a reduction in exacerbations of chronic obstructive pulmonary disease (COPD) during the coronavirus (COVID-19) pandemic, particularly hospital admissions for severe exacerbations. However, the magnitude of this reduction varies between studies.

**Method:** Electronic databases were searched from January 2020 to May 2021. Two independent reviewers screened titles and abstracts and, when necessary, full text to determine if studies met inclusion criteria. A modified version of the Newcastle-Ottawa Scale was used to assess study quality. A narrative summary of eligible studies was synthesised, and meta-analysis was conducted using a random effect model to pool the rate ratio and 95% confidence intervals (95% CI) for hospital admissions. Exacerbation reduction was compared against the COVID-19 Containment and Health Index.

**Results:** A total of 13 of 745 studies met the inclusion criteria and were included in this review, with data from nine countries. Seven studies could be included in the meta-analysis. The pooled rate ratio of hospital admissions for COPD exacerbations during the pandemic period was 0.50 (95% CI 0.42-0.58). Findings on the rate of community-treated exacerbations were inconclusive. Three studies reported a significant decrease in the incidence of respiratory viral infections compared with the pre-pandemic period. There was not a significant relationship between exacerbation reduction and the COVID-19 Containment and Health Index.

**Conclusion:** There was a 50% reduction in admissions for COPD exacerbations during the COVID-19 pandemic period, associated with a reduction in respiratory viral infections that trigger exacerbations. We provide pooled evidence supporting the potential effectiveness of COVID-19 preventive interventions in reducing the risk of admissions for COPD exacerbations.

## Introduction

Chronic obstructive pulmonary disease (COPD) affects more than 250 million people around the world and is the third leading cause of death [1]. People living with COPD are prone to acute deteriorations in health status called exacerbations [2]. The severity of exacerbations has been graded by the Global initiative for chronic Obstructive Lung Disease (GOLD) based on treatment received. Moderate exacerbations are defined as those requiring oral antibiotics and/or corticosteroids while severe exacerbations are those needing hospital admission [3].

The 2019 Coronavirus (COVID-19) pandemic has been challenging for people living with COPD. Despite the low prevalence of COPD patients in most hospital series reporting COVID-19, COPD has been associated with increased severity and mortality of COVID-19 [4, 5]. As a result, health care services were modified to reduce the risk of transmission with much contact conducted remotely [6]. In addition, policy interventions were introduced for the general public including greater hand hygiene, wearing face coverings, ‘social’ (physical) distancing, and closure of public spaces to reduce the risk of COVID-19 transmission. ‘Shielding’ or social isolation was strongly recommended in some jurisdictions for people at high-risk of poor outcomes with COVID-19, such as people living with significant COPD [7].

Most exacerbations of COPD are caused by respiratory viruses [8]. A previous meta-analysis has reported that physical interventions are able to reduce the risk of respiratory virus transmission in the general population [9]. It is therefore plausible that COVID-19 restrictions could reduce the incidence of COPD exacerbations [9]. However, shielding and other restrictions can have a negative impact on patients’ physical and mental health, and limit access to health care services [10, 11].

A number of small studies have reported an apparent reduction in the rate of COPD exacerbations during the COVID-19 pandemic, yet the precise relationship between COVID-19 preventative measures and COPD exacerbation reduction, particularly hospital admissions for severe exacerbations remain unclear. We have conducted a systematic review and meta-analysis to clarify this relationship. This is important because exacerbations have been identified as the most disruptive aspect of COPD by patients [12], current exacerbation reduction interventions are only partially effective even when deployed optimally [3], and COPD guidelines such as the GOLD document [3] and that from the European Respiratory Society/American Thoracic Society [13] make no mention of opportunities to reduce exacerbations through physical interventions to reduce the risk of acquiring respiratory viruses. A recent patient-clinician research prioritisation has identified better prevention of exacerbations as the top priority for COPD exacerbation research [14].

## Methods

We prospectively registered this systematic review and meta-analysis at PROSPERO (ID: CRD42021249522), and followed the Preferred Reporting in Systematic Reviews and Meta-Analyses (PRISMA) guidelines [15].

We used a comprehensive search strategy with assistance from a specialist librarian to retrieve relevant studies. MEDLINE and Embase via Ovid, and CINAHL were searched from 1^st^ January 2020 to 17^th^ May 2021 (see Supplementary Table S1). We also screened the reference list of selected studies. The resulting search was sent to EndNote to remove duplicates, and citations were then exported to Rayyan software for screening of title, abstract and full text by two independent reviewers.

### Data Selection

Two independent reviewers screened papers for inclusion and disagreements were resolved by discussion. We assessed the reference lists of selected papers for other eligible studies and any disagreement on included papers was resolved via discussion with a third reviewer. We included studies of COPD patients that reported non-hospitalised COPD exacerbations and/or admission rates both pre- and during the COVID-19 pandemic. The exclusion criteria comprised studies looking at pulmonary conditions other than COPD, non-full text articles, reviews, and conference abstracts.

### Quality Assessment

Two independent reviewers used a modified version of the Newcastle-Ottawa Scale (NOS) [16] to evaluate study quality. This tool included seven domains, each scored from 0 (high risk of bias) to 3 (low risk of bias). We used a mean of these domains to calculate a score between 0-3, where a higher score indicates a lower risk of bias. Discussion with a third reviewer was conducted to resolve any disagreement in the quality assessment.

### COVID-19 Containment and Health Index

We used the COVID-19 Containment and Health Index (https://ourworldindata.org/grapher/covid-containment-and-health-index) for each country and averaged the score over the reported study period. The Containment and Health Index is based on 13 measures: school closures, workplace closures, cancellation of public events, restrictions on public gatherings, closures of public transport, stay-at-home requirements, public information campaigns, restrictions on internal movements, international travel controls, testing policy, extent of contact tracing, face covering and vaccine policy. The index has a value between 0-100 and is calculated on a daily basis, a higher score indicating a stricter response. These data were extracted from Oxford Coronavirus Government Response Tracker (OxCGRT) [17], a publicly available database.

### Data analysis

We performed meta-analysis to estimate the pooled reduction in rate of COPD exacerbation hospital admissions during the COVID-19 pandemic compared with a period before COVID-19. This was completed by computing the reported rate ratio in the selected studies with 95% CI using the Metan procedure in STATA software. Studies that reported risk reduction with 95% CI were converted to rate ratio to ensure homogeneity. Studies that reported median and range rather than mean and 95% CI were converted to mean and 95% CI to standardise the meta-analysis. Corresponding authors of studies that did not report rate ratios were contacted and data were requested in the correct format. Studies that did not report a rate ratio with 95% CI and for which no reply was received from the original authors were excluded from the meta-analysis. A random-effects model in Stata/SE 16 was used to correct for between-study heterogeneity and results are presented as a forest plot. We examined between-study heterogeneity using the I^2^ statistic with corresponding p values set at 0.05. A narrative synthesis of the findings was performed considering the reduction in rate of hospitalised COPD exacerbations, rate of non-hospitalised COPD exacerbations, outcomes such as respiratory associated viral infections and which general COVID-19 measures were in place. Spearman correlation was used to examine the relationship between exacerbation reduction and the Containment and Health Index.

## Results

The preliminary search returned 745 papers of which 105 were duplicates. Title and abstract screening resulted in exclusion of 625 studies. Full-text screening was performed on the remaining 15 studies which resulted in the exclusion of another two. A total of 13 studies therefore met the inclusion/exclusion criteria and were included in the systematic review. Seven of the 13 studies presented or provided the hospital admission reduction with 95% CI and could be included in the meta-analysis (Figure 1).

**Figure 1.**
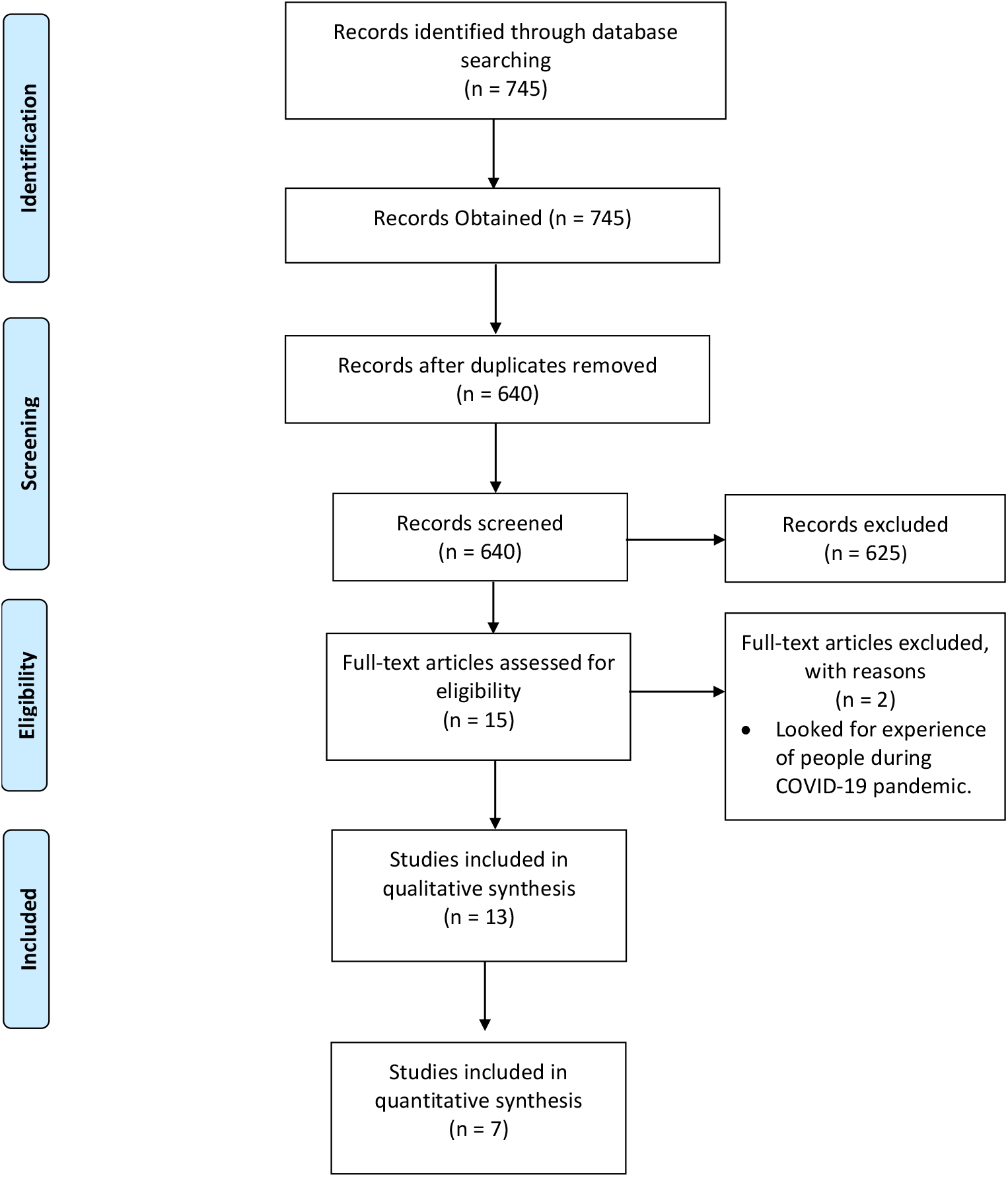
PRISMA diagram.

### Features of Included Studies

A description of the included studies is presented in Table 1. The studies included a total of 15,677 patients from nine countries (China= 2, Germany = 2, Spain= 2, United Kingdom = 2 and one each from Greece, Norway, Portugal, Republic of Korea, and Singapore). All studies were retrospective with hospital admission data between January and July 2020 considered as “during the pandemic”, compared to data from previous years (as far back as 2015). The risk of bias ranged from 0.4 to 2.5; six studies scored ≥ 2, which indicates lower risk of bias (Table S2 in the Appendix).

**Table 1.**
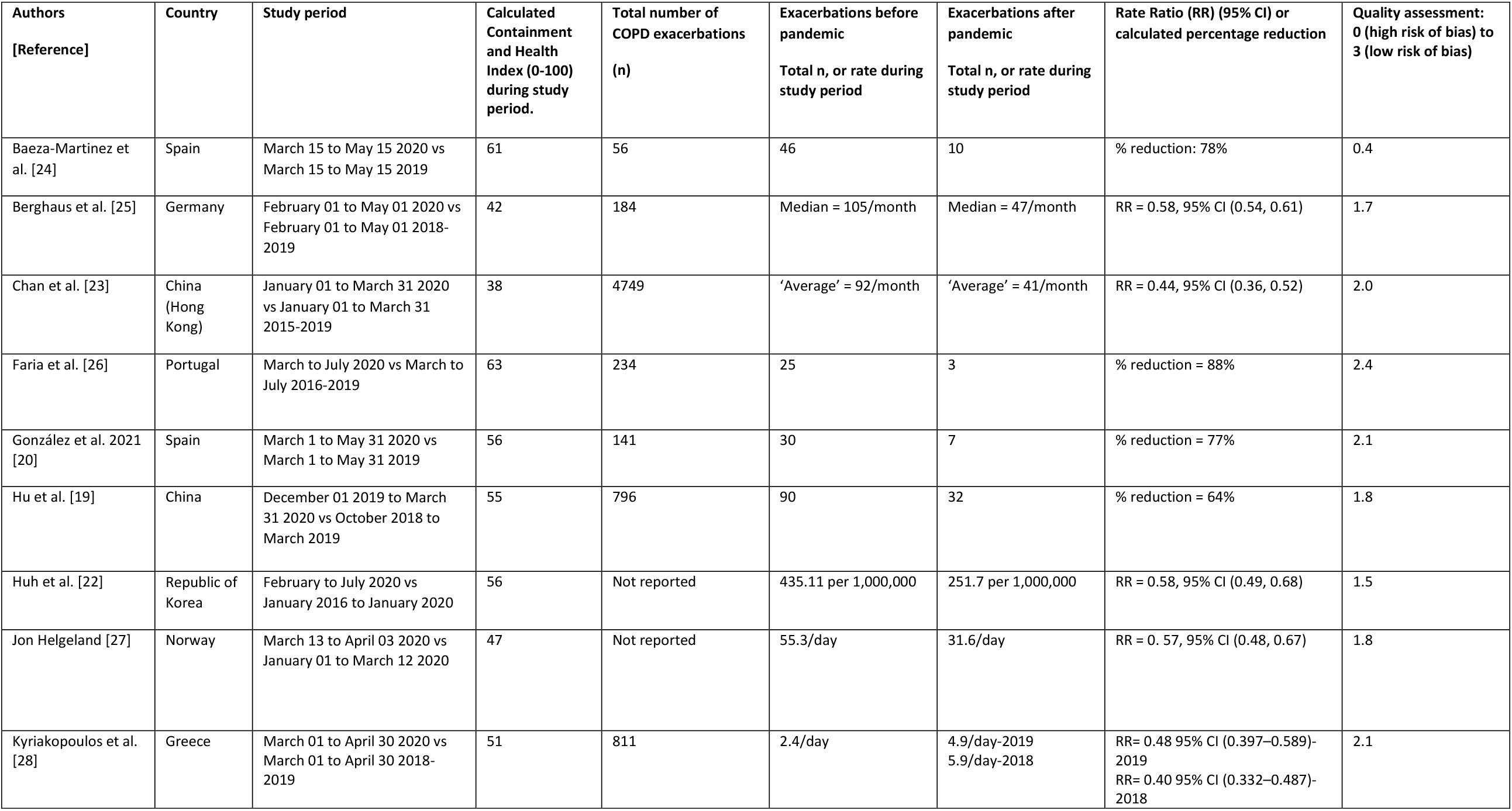
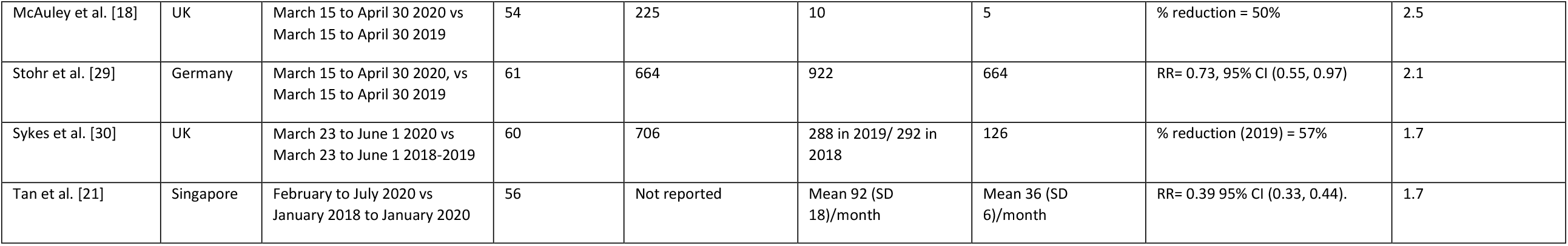
Characteristics of included studies examining reduction in hospital admissions for exacerbations of COPD.

### Reduction in hospital admissions for COPD Exacerbations

All studies reported a reduction in hospital admissions for COPD exacerbations in association with the COVID-19 pandemic, with the calculated percentage reduction varying between 27% and 88%. The highest and lowest percentage reduction were found in Portugal and Germany respectively (Figure 2). The pooled rate ratio for the reduction of hospital admissions for COPD exacerbation during COVID-19 pandemic compared to the pre-COVID-19 period was 0.50 (95% CI, 0.42-0.58). The measure of between-studies heterogeneity (I^2^) was 86.8%, p <0.001 (Figure 3).

**Figure 2.**
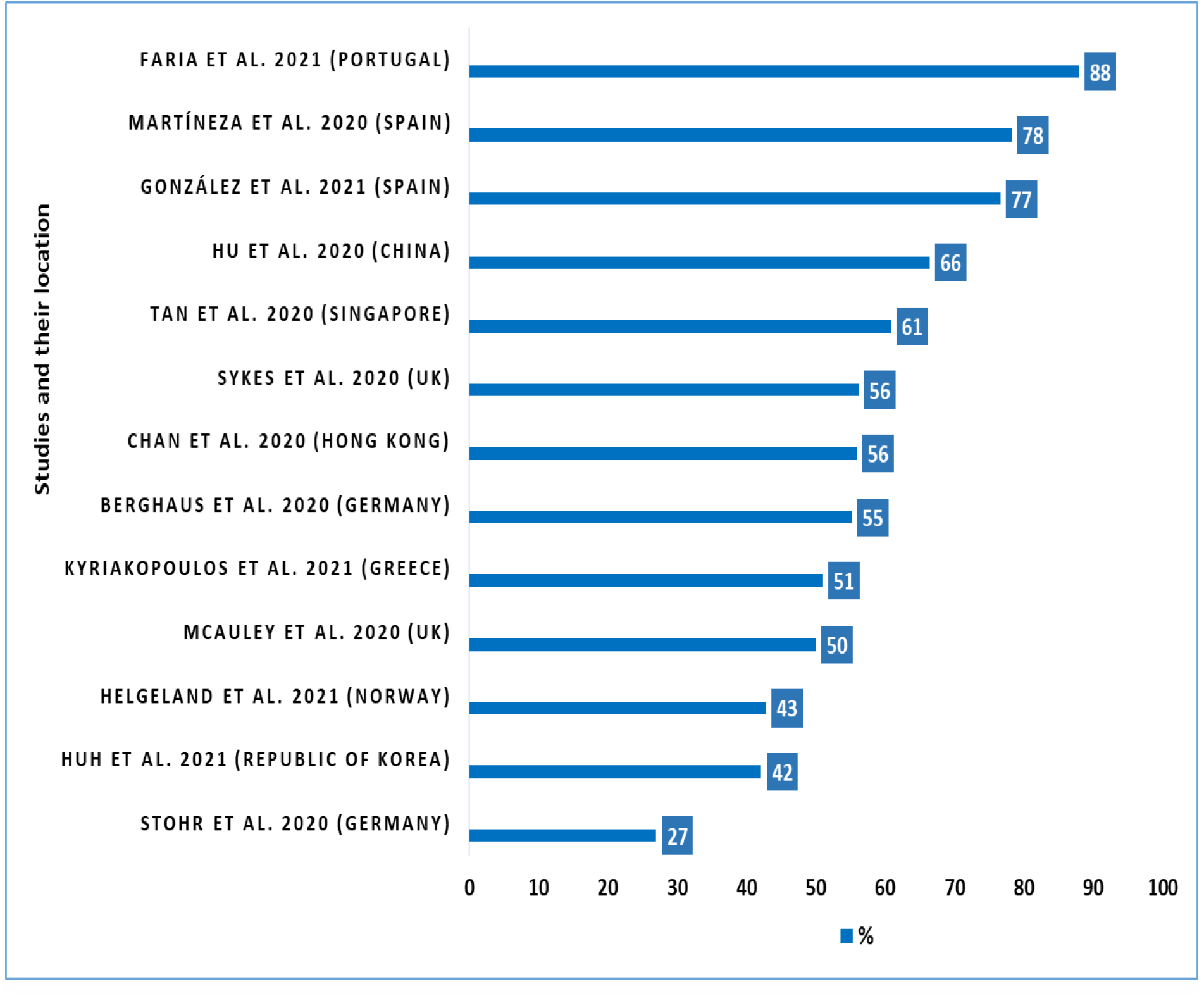
Calculated percentage reduction in COPD exacerbations during the COVID-19 pandemic across 13 studies.

**Figure 3.**
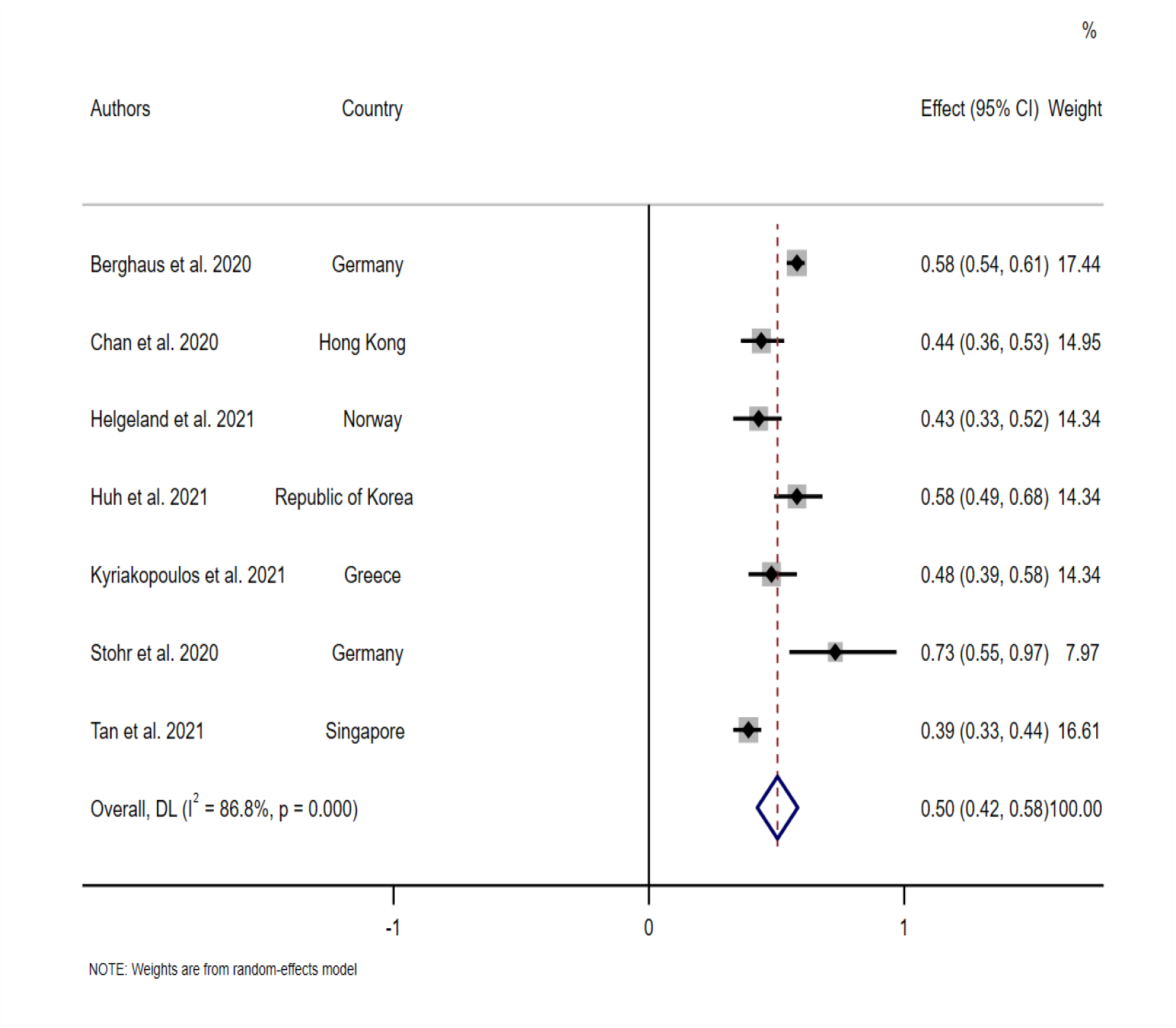
Pooled rate ratio of hospitalised COPD exacerbations in the post-compared to the pre-COVID-19 pandemic period.

### Reduction in community-treated COPD Exacerbations

Only three studies investigated COPD exacerbations managed in the community [18-20]. McAuley et al. [18] reported an increase in community managed exacerbations of 38%, in which 121 exacerbations were observed during the pandemic (2020) compared to 88 exacerbations in the pre-pandemic period (2019). In contrast, Hu et al. [19] reported a 39% reduction in exacerbations during the pandemic compared to the pre-pandemic period (97 events vs 160 events) respectively. Moreover, they reported 80% (392) of their patients remained in a stable condition. In Spain, there was a 55% reduction in non-hospitalised COPD exacerbations (32 events vs 72 events) compared to the pre-pandemic period [20].

### Reduction in respiratory viral infections

Three studies (all from Asia: China (Hong Kong), Republic of Korea and Singapore) assessed the prevalence of respiratory viral infections in association with exacerbations [21-23]. Despite increased testing for respiratory viruses during the pandemic (pre-pandemic: 60% vs pandemic: 98%), Tan et al. [21] reported the incidence rate ratio of positive viral infections in exacerbations admissions was reduced at 0.35, 95% CI (0.22 to 0.53, p<0.001). Huh et al. [22] assessed hospitalisations due to pneumonia or influenza in those with pre-existing COPD and found a reduction in both conditions during the pandemic compared to the pre-pandemic period, rate ratio (95% CI): 0.54 (0.53-0.55) and 0.27 (0.24-0.30) respectively. In Hong Kong, a reduction in the number of influenza A or B viruses in people with COPD was observed during the pandemic compared with pre-pandemic period (monthly average: 1888 vs 3958 respectively) [23]. The reduction was associated with a decrease in exacerbations.

### COVID-19 transmission reduction measures

The COVID-19 Containment and Health Index data are reported in Table 1. Although, in general, there was the suggestion that more stringent containment and health may be associated with a greater reduction in exacerbations, this did not reach statistical significance (rho= 0.34, p= 0.26).

## Discussion

This systematic review and meta-analysis reports a 50% reduction in hospital admissions for COPD exacerbations during the COVID-19 pandemic compared to pre-pandemic times. Studies from nine countries consistently report a reduction. In a subset of included studies, there was a reduction in respiratory viral infections suggesting that a major mechanism for reduced exacerbations is likely reduced respiratory virus transmission. Data for community-treated exacerbations were inconclusive and need further investigation. This data is the first to provide pooled evidence supporting the potential effectiveness of COVID-19 preventative interventions in reducing the transmission of respiratory viruses, ultimately reducing admissions for COPD exacerbations – a major area of unmet clinical need and research priority [14].

The aim of this systematic review was to define the relationship between COVID-19 related preventive measures and reduction in COPD exacerbations, especially the most severe exacerbations that result in hospital admission. The reduction in admissions for COPD exacerbation during the COVID-19 pandemic indicates the potential benefit of continued implementation of physical interventions in reducing viral transmission. The reported reductions ranged from 27% to 88%, and 10 of the 13 included studies reported a ≥50% reduction in hospitalised COPD exacerbation admissions [18-21, 23-26, 28, 30]. There was not a statistically significant relationship between the COVID-19 Containment and Health Index and magnitude of exacerbation reduction. Nevertheless, the reduction in hospitalisation is greater than ever achieved with existing optimisation of COPD care [3]. Others have noted a reduction in hospitalised asthma exacerbations by 36% [31] and 79% [28].

There are several explanations for our findings. We have summarised these in a directed acyclic graph (DAG, Figure 4) [32]. The reduction in respiratory viruses associated with residual COPD exacerbations (30) suggests that a major mechanism driving exacerbation reduction is a reduction in acquisition of respiratory viruses, likely through hand hygiene, face coverings and physical distancing in the general population, and shielding in those with COPD. However, there are alternative explanations. The absence of a correlation between COVID-19 Containment and Health Index and exacerbation reduction may suggest that individual behaviour in people living with COPD is more impactful than the changes instituted at societal level. Anxiety in association with COVID-19 might have resulted in an increase in medication adherence [18, 30]. Air pollutants add to the risk of COPD exacerbations, although the effect is less strong than that for respiratory viruses [33], and emissions of pollutants such as nitrogen dioxide (NO_2_) [34] and indeed smoking [35] were reported to be significantly reduced during the pandemic. More concerningly, the reduction in COPD admissions could also reflect decreased presentation at hospitals, perhaps through fear and limited access. However, Davies et al. used cause-specific mortality data in asthma to explore this and found that a reduction in admissions was not associated with an increase in mortality, suggesting a real reduction in the incidence of respiratory exacerbations [31]. In Germany, COPD hospitalisations were compared with admissions for other conditions including myocardial infarction and stroke and the reduction in COPD admissions was greater than that observed in other conditions [25]. This also suggests that the reduction in COPD exacerbations in association with COVID-19 restrictions is indeed a real reduction rather than under-representation of COPD patients attending hospital services.

**Figure 4.**
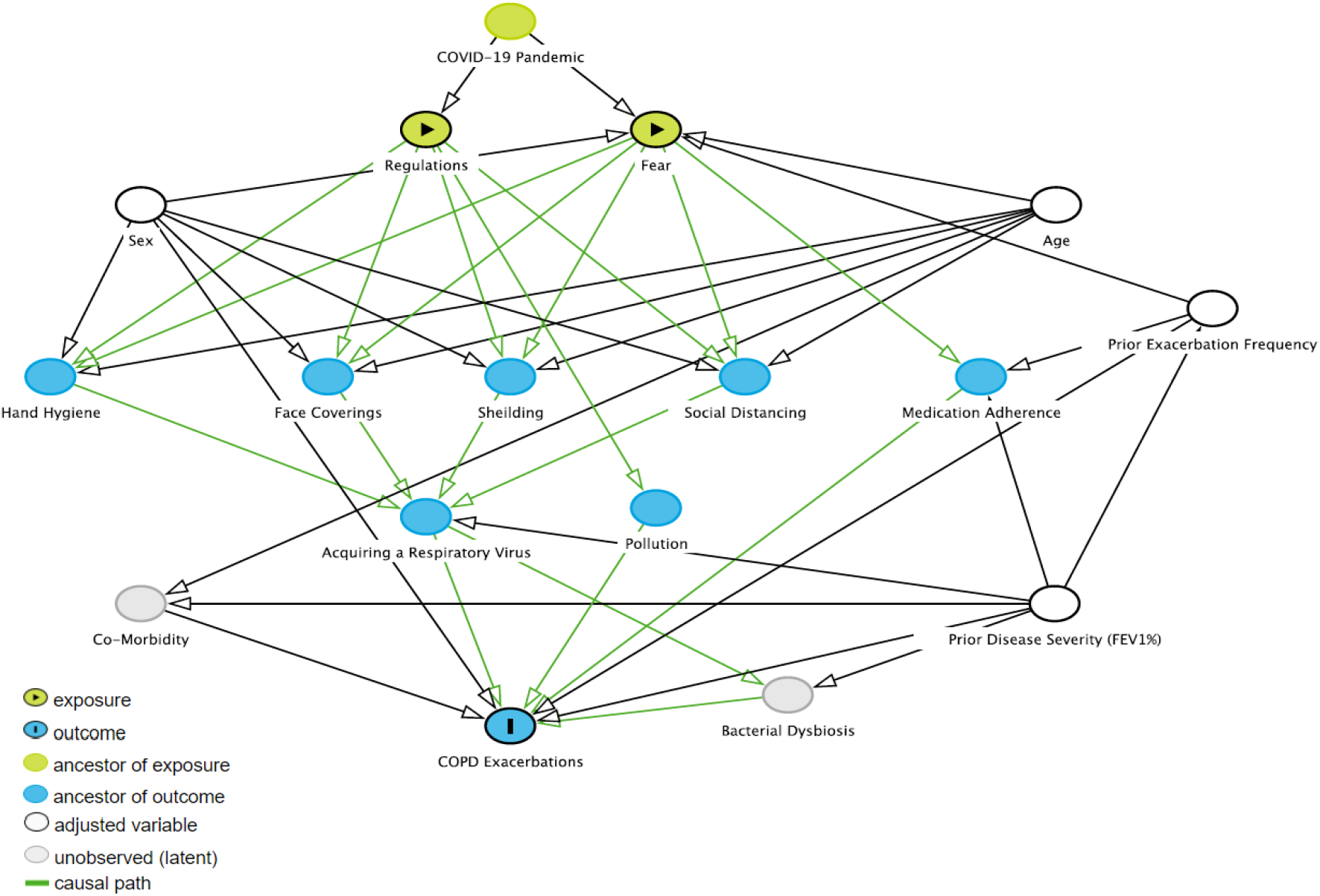
Directed acyclic graph describing relationships between the COVID-19 pandemic and a reduction in COPD exacerbations. We first considered factors known to be associated with COPD exacerbation risk (the outcome). We then considered respiratory virus infection control interventions developed during the pandemic (‘regulations’), and other variables associated with the pandemic that might associate with exacerbation risk: fear of coronavirus. We therefore considered regulation and fear to be the exposures, both arising from the COVID-19 pandemic. The DAG suggests that age, sex, prior exacerbation frequency and prior disease severity (FEV_1_ % predicted) are potential confounders.

In general, COVID-19 restrictions have had a negative impact on those living with COPD. Two studies from the UK assessed patients’ experience during the pandemic and how it affected disease control and mental health [18, 30]. Sykes et al. [30] reported 32% (16/50) of interviewed patients had worsening of their disease control, 46% (23/50) reported a reduction in daily exercise and 48% (24/50) stated that COVID-19 negatively affected their mental health. The use of bronchodilator inhalers was increased in 48% (24/50) of patients. Similarly, McAuley et al. [18] reported an increase in anxiety by 58% (92/160), and inhaler adherence, whilst physical activity was lower such that only 16% (26/160) kept the same level of physical activity. Data obtained from a survey of 100 Spanish COPD patients showed that 64% completely complied with lockdown, 90% had medical visits or tests cancelled and 37% were afraid of death [36].

Despite the high burden of COPD exacerbations on society, infection control measures against respiratory viruses associated with exacerbations are not currently included in COPD clinical management guidelines. Guidelines primarily focus on optimisation of pharmacological and non-pharmacological interventions, and recommendations to actively reduce viral transmission are not provided. A Cochrane review has highlighted the efficacy of simple, low-cost physical interventions such as hand hygiene and face coverings to reduce respiratory virus transmission in the general population, although endorsing routine use was noted to be potentially difficult in a non-epidemic period [9]. A recent study assessed the acceptability to people living with respiratory disease of continuing such physical interventions to prevent exacerbations of airways diseases beyond the pandemic [37]. People living with COPD, asthma and bronchiectasis were supportive of some on-going infection control interventions and the authors recommended that such measures be considered in future guidelines [37]. Acceptance of restrictions varied by age and sex (and these influences are included in our DAG). We strongly advocate that the reduction in the rate of hospital admissions for COPD exacerbation during the COVID-19 pandemic and the positive attitudes of people living with airways diseases toward maintaining adoption of physical interventions indicates a need to reassess infection control measures in future COPD guidelines. Including such measures could potentially also reduce the high burden of hospital re-admissions for COPD exacerbations [38, 39]. Novel strategies to prevent COPD exacerbations have been identified as a top priority by patients and clinicians in a recent research prioritisation exercise [14].

These analyses have strengths and limitations. This is the first systematic analysis of the impact of COVID-19 pandemic restrictions on COPD exacerbations, especially the most severe COPD exacerbations requiring hospital admission. The results from nine countries were consistent, and we were able to pool data to estimate the weighted rate ratio, considering variation between studies. There was heterogeneity between studies in location and follow-up time while some studies did not report the rate ratio with 95% CI and could not be included in the meta-analysis. Restrictions within individual countries changed with time and we were not able to confirm that jurisdictions with the greatest restrictions saw the greatest reduction in exacerbations. Our DAG permits identification of confounders in the relationship between the coronavirus pandemic and reduced admissions to hospital for COPD exacerbations. Key confounders are age, sex, prior exacerbation frequency and prior disease severity. These confounders have not been accounted for in studies published to date.

In conclusion, this systematic analysis and meta-analysis reports a 50% reduction in hospital admissions for COPD exacerbations during the COVID-19 pandemic, most likely primarily through reduction in transmission of respiratory viruses. Data for community treated exacerbations were inconclusive and this requires further examination. This work has important implications for future decision-making regarding the prevention of COPD exacerbations. Future guidelines should include recommendations on respiratory virus infection control measures to reduce the burden of COPD exacerbations beyond the pandemic period.

## Data Availability

All data included in the manuscript.

## Appendix

**Table S1:**
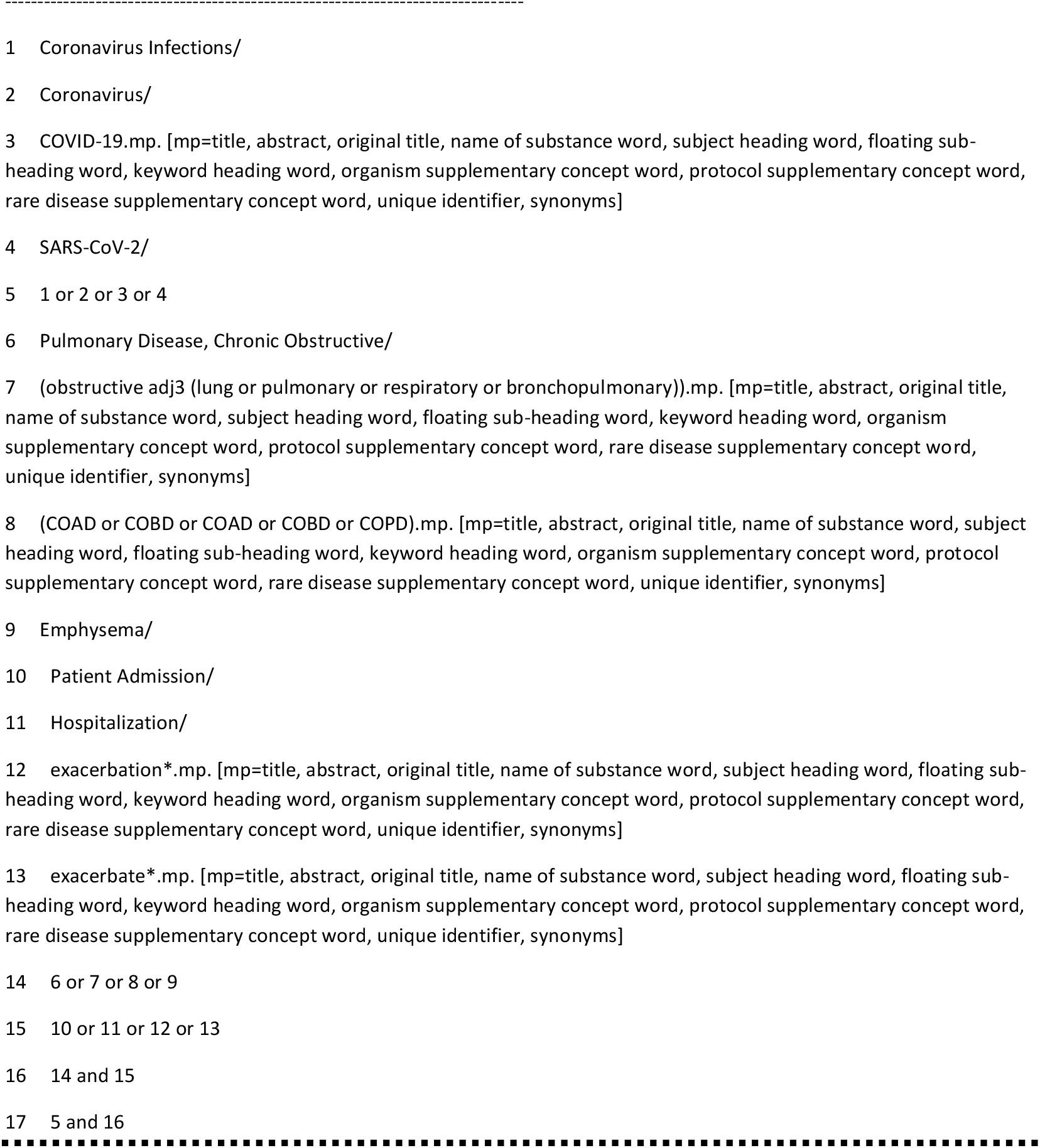
Medline search strategy. Database: Ovid MEDLINE(R) and Epub Ahead of Print, In-Process, In-Data-Review & Other Non-Indexed Citations and Daily Search Strategy:

**Table S2.** Quality assessment. Details of the quality of Cohort studies

| First author | Population representative | Sample size adequate | Confounders | Statistical analysis | Missing data | Methodology of the outcome | Objective assessment | OVERALL (0-3, higher score = lower risk of bias) |
| --- | --- | --- | --- | --- | --- | --- | --- | --- |
| Baeza-Martínez et al. 2020 | 1 | 0 | 0 | 0 | 2 | 0 | 0 | 0.4 |
| Berghaus et al. 2020 | 2 | 1 | 1 | 2 | 1 | 3 | 2 | 1.7 |
| Chan et al. 2020 | 3 | 1 | 1 | 3 | 1 | 3 | 2 | 2 |
| Faria et al. 2021 | 3 | 1 | 2 | 3 | 2 | 3 | 3 | 2.4 |
| González et al. 2021 | 3 | 2 | 1 | 3 | 1 | 2 | 3 | 2.1 |
| Helgeland et al. 2021 | 2 | 0 | 1 | 3 | 1 | 3 | 3 | 1.8 |
| Hu et al. 2020 | 2 | 2 | 1 | 3 | 1 | 3 | 1 | 1.8 |
| Huh et al. 2021 | 2 | 1 | 1 | 2 | 1 | 2 | 2 | 1.5 |
| Kyriakopoulos et al. 2021 | 2 | 1 | 2 | 3 | 2 | 2 | 3 | 2.1 |
| McAuley et al. 2020 | 3 | 2 | 1 | 3 | 3 | 3 | 3 | 2.5 |
| Stohr et al. 2020 | 2 | 3 | 1 | 3 | 1 | 2 | 3 | 2.1 |
| Sykes et al. 2020 | 2 | 1 | 0 | 3 | 1 | 3 | 2 | 1.7 |
| Tan et al. 2020 | 2 | 1 | 1 | 2 | 1 | 2 | 3 | 1.7 |

0 = definitely no (high risk of bias); 1 = mostly no; 2 = Mostly yes; 3 = definitely yes (low risk of bias)

## Notes

### Competing Interest Statement

The authors have declared no competing interest.

### Funding Statement

No fund.

## References

1. WHO. Chronic obstructive pulmonary disease (COPD) 2020. Available from: https://www.who.int/news-room/fact-sheets/detail/chronic-obstructive-pulmonary-disease-(copd)

2. Hurst JR, Vestbo J, Anzueto A, Locantore N, Müllerova H, Tal-Singer R, et al. Susceptibility to exacerbation in chronic obstructive pulmonary disease. The New England journal of medicine. 2010;363(12):1128-38. Epub 2010/09/17. doi: 10.1056/NEJMoa0909883. PubMed PMID: 20843247.

3. Global Initiative for Chronic Obstructive Lung Disease. Global Strategy for the Diagnosis, Management, and Prevention of Chronic Obstructive Pulmonary Disease. 2021 [cited 2021 01/03/2021]. Available from: https://goldcopd.org/wp-content/uploads/2019/11/GOLD-2020-POCKET-GUIDE-FINAL-pgsized-wms.pdf.

4. Alqahtani JS, Oyelade T, Aldhahir AM, Alghamdi SM, Almehmadi M, Alqahtani AS, et al. Prevalence, Severity and Mortality associated with COPD and Smoking in patients with COVID-19: A Rapid Systematic Review and Meta-Analysis. PloS one. 2020;15(5):e0233147–e. doi: 10.1371/journal.pone.0233147. PubMed PMID: 32392262.

5. Gerayeli FV, Milne S, Cheung C, Li X, Yang CWT, Tam A, et al. COPD and the risk of poor outcomes in COVID-19: A systematic review and meta-analysis. EClinicalMedicine. 2021;33:100789. Epub 2021/03/25. doi: 10.1016/j.eclinm.2021.100789. PubMed PMID: 33758801; PubMed Central PMCID: PMCPMC7971471.

6. WHO. Home care for patients with suspected or confirmed COVID-19 and management of their contacts. [cited 23/04/2021]. Available from: https://www.who.int/publications/i/item/home-care-for-patients-with-suspected-novel-coronavirus-(ncov)-infection-presenting-with-mild-symptoms-and-management-of-contacts.

7. Guidance on shielding and protecting people who are clinically extremely vulnerable from COVID-19. Available from: https://www.gov.uk/government/publications/guidance-on-shielding-and-protecting-extremely-vulnerable-persons-from-covid-19/guidance-on-shielding-and-protecting-extremely-vulnerable-persons-from-covid-19.

8. Hewitt R, Farne H, Ritchie A, Luke E, Johnston SL, Mallia P. The role of viral infections in exacerbations of chronic obstructive pulmonary disease and asthma. Ther Adv Respir Dis. 2016;10(2):158-74. Epub 2015/11/28. doi: 10.1177/1753465815618113. PubMed PMID: 26611907; PubMed Central PMCID: PMCPMC5933560.

9. Jefferson T, Del Mar CB, Dooley L, Ferroni E, Al-Ansary LA, Bawazeer GA, et al. Physical interventions to interrupt or reduce the spread of respiratory viruses. The Cochrane database of systematic reviews. 2011;2011(7):Cd006207. Epub 2011/07/08. doi: 10.1002/14651858.CD006207.pub4. PubMed PMID: 21735402; PubMed Central PMCID: PMCPMC6993921.

10. Pfefferbaum B, North CS. Mental Health and the Covid-19 Pandemic. New England Journal of Medicine. 2020;383(6):510–2. doi: 10.1056/NEJMp2008017.

11. Wu F, Burt J, Chowdhury T, Fitzpatrick R, Martin G, van der Scheer JW, et al. Specialty COPD care during COVID-19: patient and clinician perspectives on remote delivery. BMJ open respiratory research. 2021;8(1):e000817. doi: 10.1136/bmjresp-2020-000817. PubMed PMID: 33414261.

12. Zhang Y, Morgan RL, Alonso-Coello P, Wiercioch W, Bała MM, Jaeschke RR, et al. A systematic review of how patients value COPD outcomes. European Respiratory Journal. 2018;52(1):1800222. doi: 10.1183/13993003.00222-2018.

13. Wedzicha JAEC-C, Miravitlles M, Hurst JR, Calverley PM, Albert RK, Anzueto A, et al. Management of COPD exacerbations: a European Respiratory Society/American Thoracic Society guideline. Eur Respir J. 2017;49(3). Epub 2017/03/17. doi: 10.1183/13993003.00791-2016. PubMed PMID: 28298398.

14. Alqahtani JS, Aquilina J, Bafadhel M, Bolton CE, Burgoyne T, Holmes S, et al. Research priorities for exacerbations of COPD. The Lancet Respiratory Medicine. doi: 10.1016/S2213-2600(21)00227-7.

15. Moher D, Shamseer L, Clarke M, Ghersi D, Liberati A, Petticrew M, et al. Preferred reporting items for systematic review and meta-analysis protocols (PRISMA-P) 2015 statement. Syst Rev. 2015;4:1. Epub 2015/01/03. doi: 10.1186/2046-4053-4-1. PubMed PMID: 25554246; PubMed Central PMCID: PMCPMC4320440.

16. GA Wells BS, D O’Connell, J Peterson, V Welch, M Losos, P Tugwell. The Newcastle-Ottawa Scale (NOS) for assessing the quality of nonrandomised studies in meta-analyses [cited 2019 23/01]. Available from: http://www.ohri.ca/programs/clinical_epidemiology/oxford.asp

17. Hale T, Angrist N, Goldszmidt R, Kira B, Petherick A, Phillips T, et al. A global panel database of pandemic policies (Oxford COVID-19 Government Response Tracker). Nature Human Behaviour. 2021;5(4):529–38. doi: 10.1038/s41562-021-01079-8.

18. McAuley H, Hadley K, Elneima O, Brightling CE, Evans RA, Steiner MC, et al. COPD in the time of COVID-19: an analysis of acute exacerbations and reported behavioural changes in patients with COPD. Erj Open Research. 2021;7(1). PubMed PMID: 33527075.

19. Hu W, Dong M, Xiong M, Zhao D, Zhao Y, Wang M, et al. Clinical Courses and Outcomes of Patients with Chronic Obstructive Pulmonary Disease During the COVID-19 Epidemic in Hubei, China. International Journal of Copd. 15:2237-48. PubMed PMID: 33061341.

20. González J, Moncusí-Moix A, Benitez ID, Santisteve S, Monge A, Fontiveros MA, et al. Clinical consequences of COVID-19 lockdown in patients with COPD: results of a pre-post study in Spain. Chest. 2021.

21. Tan JY, Conceicao EP, Wee LE, Sim XYJ, Venkatachalam I. COVID-19 public health measures: a reduction in hospital admissions for COPD exacerbations. Thorax. 2020;03:03. PubMed PMID: 33273024.

22. Huh K, Kim YE, Ji W, Kim DW, Lee EJ, Kim JH et al. Decrease in hospital admissions for respiratory diseases during the COVID-19 pandemic: a nationwide claims study. Thorax. 2021 Mar 29:thoraxjnl-2020-216526. doi: 10.1136/thoraxjnl-2020-216526. Epub ahead of print. PMID: 33782081; PMCID: PMC8011422.

23. Chan KPF, Ma TF, Kwok WC, Leung JKC, Chiang KY, Ho JCM, et al. Significant reduction in hospital admissions for acute exacerbation of chronic obstructive pulmonary disease in Hong Kong during coronavirus disease 2019 pandemic. Respiratory Medicine. 171:106085. PubMed PMID: 32917356.

24. Baeza-Martinez C, Zamora-Molina L, Olea-Soto J, Soler-Sempere MJ, Garcia-Pachon E. Reduction in Hospital Admissions for COPD Exacerbation During the Covid-19 Pandemic. Open Respiratory Archives. 2020;2(3):201-2. PubMed PMID: 2007104067.

25. Berghaus TM, Karschnia P, Haberl S, Schwaiblmair M. Disproportionate decline in admissions for exacerbated COPD during the COVID-19 pandemic. Respiratory Medicine.106120. PubMed PMID: 32839072.

26. Faria N, Costa MI, Gomes J, Sucena M. Reduction of Severe Exacerbations of COPD during COVID-19 Pandemic in Portugal: A Protective Role of Face Masks? Copd: Journal of Chronic Obstructive Pulmonary Disease.1-9. PubMed PMID: 33764237.

27. Helgeland J, Telle KE, Grosland M, Huseby BM, Haberg S, Lindman ASE. Admissions to Norwegian Hospitals during the COVID-19 Pandemic. Scandinavian journal of public health. 2021:14034948211000813. PubMed PMID: 634661423.

28. Kyriakopoulos C, Gogali A, Exarchos K, Potonos D, Tatsis K, Apollonatou V, et al. Reduction in Hospitalizations for Respiratory Diseases during the First COVID-19 Wave in Greece. Respiration; international review of thoracic diseases. 2021:1–6. Epub 2021/04/08. doi: 10.1159/000515323. PubMed PMID: 33827103; PubMed Central PMCID: PMCPMC8089411.

29. Stohr E, Aksoy A, Campbell M, Al Zaidi M, Ozturk C, Vorloeper J, et al. Hospital admissions during Covid-19 lock-down in Germany: Differences in discretionary and unavoidable cardiovascular events. PLoS ONE. 15(11):e0242653. PubMed PMID: 33216804.

30. Sykes DL, Faruqi S, Holdsworth L, Crooks MG. Impact of COVID-19 on COPD and asthma admissions, and the pandemic from a patient’s perspective. Erj Open Research. 2021;7(1). PubMed PMID: 33575313.

31. Davies GA, Alsallakh MA, Sivakumaran S, Vasileiou E, Lyons RA, Robertson C, et al. Impact of COVID-19 lockdown on emergency asthma admissions and deaths: national interrupted time series analyses for Scotland and Wales. Thorax. 2021. doi: 10.1136/thoraxjnl-2020-216380. PubMed PMID: 33782079.

32. Textor J, van der Zander B, Gilthorpe MS, Liskiewicz M, Ellison GT. Robust causal inference using directed acyclic graphs: the R package ‘dagitty’. Int J Epidemiol. 2016;45(6):1887–94. Epub 2017/01/17. doi: 10.1093/ije/dyw341. PubMed PMID: 28089956.

33. Anderson JO, Thundiyil JG, Stolbach A. Clearing the air: a review of the effects of particulate matter air pollution on human health. Journal of medical toxicology : official journal of the American College of Medical Toxicology. 2012;8(2):166–75. Epub 2011/12/24. doi: 10.1007/s13181-011-0203-1. PubMed PMID: 22194192; PubMed Central PMCID: PMCPMC3550231.

34. The Air Quality Expert Group to the Department for Environment, Food and Rural Affairs. Estimation of changes in air pollution emissions, concentrations and exposure during the COVID-19 outbreak in the UK. rapid evidence review– June 2020. Available from: https://uk-air.defra.gov.uk/assets/documents/reports/cat09/2007010844_Estimation_of_Changes_in_Air_Pollution_During_COVID-19_outbreak_in_the_UK.pdf.

35. Action on Smoking and Health. A million people have stopped smoking since the COVID pandemic hit Britain. . Available from: https://ash.org.uk/media-and-news/press-releases-media-and-news/pandemicmillion/.

36. Pleguezuelos E, Del Carmen A, Moreno E, Ortega P, Vila X, Ovejero L, et al. The Experience of COPD Patients in Lockdown Due to the COVID-19 Pandemic. Int J Chron Obstruct Pulmon Dis. 2020;15:2621–7. Epub 2020/10/31. doi: 10.2147/copd.S268421. PubMed PMID: 33122900; PubMed Central PMCID: PMCPMC7591044.

37. Hurst JR, Cumella A, Niklewicz CN, Philip KEJ, Singh V, Hopkinson NS. Long-Term Acceptability of Hygiene, Face Covering, and Social Distancing Interventions to Prevent Exacerbations in people living with Airways Diseases. medRxiv. 2021:2021.04.09.21255189. doi: 10.1101/2021.04.09.21255189.

38. Alqahtani JS, Njoku CM, Bereznicki B, Wimmer BC, Peterson GM, Kinsman L, et al. Risk factors for all-cause hospital readmission following exacerbation of COPD: a systematic review and meta-analysis. European Respiratory Review. 2020;29(156):190166. doi: 10.1183/16000617.0166-2019.

39. Njoku CM, Alqahtani JS, Wimmer BC, Peterson GM, Kinsman L, Hurst JR, et al. Risk factors and associated outcomes of hospital readmission in COPD: A systematic review. Respir Med. 2020;173:105988. Epub 2020/11/17. doi: 10.1016/j.rmed.2020.105988. PubMed PMID: 33190738.

